# The Futures of the Pandemic in the USA: A Timed Intervention Model

**DOI:** 10.1101/2020.08.23.20180174

**Authors:** Gary D. Hachtel, John D. Stack, Jordan A. Hachtel

## Abstract

We propose a novel **Timed Intervention** *S, P, E, I, Q, R, D* model for projecting the possible futures of the COVID-19 pandemic in the USA. The proposed model introduces a series of timed interventions that can account for the influence of real time changes in government policy and social norms. We consider three separate types of interventions:

Protective interventions. Where population moves from susceptible to protected corresponding to mask mandates, stay-at-home orders and/or social distancing.
Release interventions. Where population moves from protected to susceptible corresponding to social distancing mandates and practices being lifted by policy or pandemic fatigue.
Vaccination interventions. Where population moves from susceptible, protected, and exposed to recovered (meaning immune) corresponding to the mass immunization of the U.S. Population.

By treating the pandemic with timed interventions, we are able to model the pandemic extremely effectively, as well as directly predicting of the course of the pandemic under differing sets of intervention schedules. We show that without prompt effective protective/vaccination interventions the pandemic will extend all the way into 2022 and result in many millions of deaths in the U.S.^†^

**Copyright Notice:** This manuscript has been authored by UT-Battelle, LLC under Contract No. DE-AC05-00OR22725 with the U.S. Department of Energy. The United States Government retains and the publisher, by accepting the article for publication, acknowledges that the United States Government retains a non-exclusive, paid-up, irrevocable, worldwide license to publish or reproduce the published form of this manuscript, or allow others to do so, for United States Government purposes. The Department of Energy will provide public access to these results of federally sponsored research in accordance with the DOE Public Access Plan (http://energy.gov/downloads/doe-public-access-plan).

## 1 Introduction

The pandemic has led to increased world interest in epidemiological modeling and prediction research, much of which focuses on extensions of the dynamic SIR model of Reference [1], developed for modeling the Spanish Flu pandemic of 1917-18, which was exacerbated by the super-spreading of the trench warfare of WWI. Some of the recent relevant academic research has been reported in References [2, 3, 4, 5]. Here we propose a new model which can, by parameter assignment, be reduced to the model of [2], which appeared in February, 2020. Like the model of Reference [2], the model proposed here partitions the total population into 7 states (or “compartments”). Unlike the model of [5], no explicit delays are included in the formulation. Rather, delays arise as an inherent property of the parameter assignment, and the structure of the state equations in Equation 1 below.

The growth of populations as diverse as yeast cultures and national populations was studied in a very general setting in [6]. In that work, Pearl showed that the logistic function provided a quantitative model for the growth of various populations. Similarly Rappole[7] used logistic functions to model habitat control of avian population, again employing logistic modeling. In our study, it is found that logistic-like functions model sink states like the recovered and deceased states *R* and *D*, but that Gaussian-like functions are needed to model the non-sink states exposed, infected, and quarantined states *E, I*, and *Q*, since these states all exhibit extrema in their time evolution. The timed interventions directly control the waveforms of the susceptible and protected states *S*, and *P*, which would normally be modeled by logistic-like functions but have a piecewise waveform in our model that is only logistic-like between successive interventions.

Epidemics are not rare but are frequent visitors to the world stage, like earthquakes and category 5 hurricanes. As a result, there is an inherent need for established and validated long term forecasting of epidemics, to help guide policy making decisions and establish cost-benefit analyses [8, 9]. This is all the more critical during the current COVID-19 crisis.

### 1.1 The COVID-19 Pandemic in the USA

Figure 1 demonstrates the primary motivation for developing the proposed timed intervention model. These deaths per day data show the alarming rise observed between day 50 and day 78, as well as the equally alarming second and third upsurges which began around days 160 and 265.

**Figure 1:**
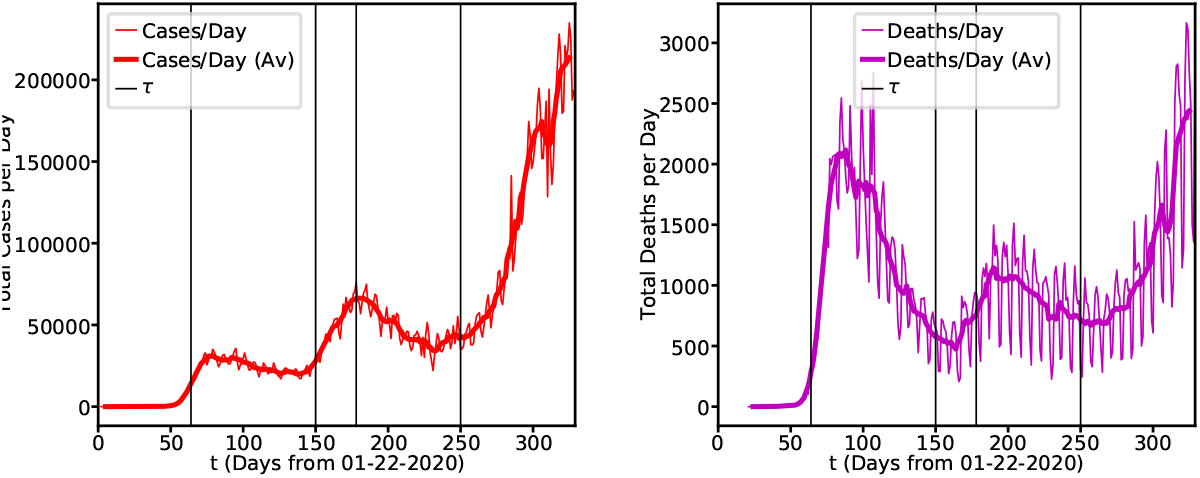
Smoothed Deaths per day source data

The smoothed (7-day rolling average) waveform appears to have 5 distinct phases. The first, for 0 < *t* ≤ 64, could be called a “preliminary spread” phase. In this phase the outbreak established itself in the state of Washington. However, we see that around day 64 the preliminary spread began to slow, reaching a peak in cases per day at around day 78 and then beginning to decrease. Day 64 would be March 26^th^, 2020 which was approximately the time where stay-at-home orders were beginning to be issued nationwide. Thus, from 64 < *t* ≤ 150, the number of new cases per day stabilizes and starts to decrease resulting in a significant decrease in deaths per day, we call this a “protective” phase. However, we see that at around day 150 the cases begin to trend sharply upwards again, which likely corresponds to the lifting of the stay-at-home orders and the general population relaxing their commitment to social distancing guidelines, we call this a “release” phase. It can be seen clearly that the release phase lasts from 150 < *t* ≤ 178, where another protective phase begins which goes from 178 < *t* ≤ 250. The most alarming trend is that since day 250 the U.S. has been in an extended release phase, with cases per day and deaths per day spiking to unprecedented levels. Clearly another protective phase is needed.

The classical predator-prey models of the Lotka-Volterra equations that first appeared in [1] do not show such articulated multi-phase behavior. Similarly, more modern work on pandemic modeling [2] also does not show such behavior, necessitating the development of a new model which does.

### 1.2 Proposed Timed Intervention Model

Our dynamical pandemic model is represented by the following state equations

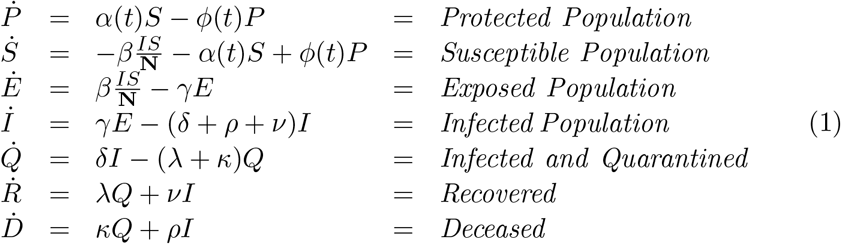

It should be mentioned at the outset that state *P* of this model does not strictly correspond to the state of the same name in [2], where *P* stood for an auto-immune population. In the proposed model *P* stands for a protected, i.e. “sheltered in place” or “social distancing”, population and any immune population is lumped into the *R* state. A graph showing the directional flow of population is given in the state transition diagram of Figure 2. Increments of population flow systematically through this graph like clockwork, in which the time step is 1 day. The lack of a *E* to *S* arrow in this figure indicates that, once exposed, members of the exposed population cannot rejoin the ranks of the susceptible population. Similarly, once infected, members of the infected population cannot rejoin the members of the exposed population. This implies that individuals *can not be re-infected*. The only feedback loop is from *S* to *P* and back.

**Figure 2:**
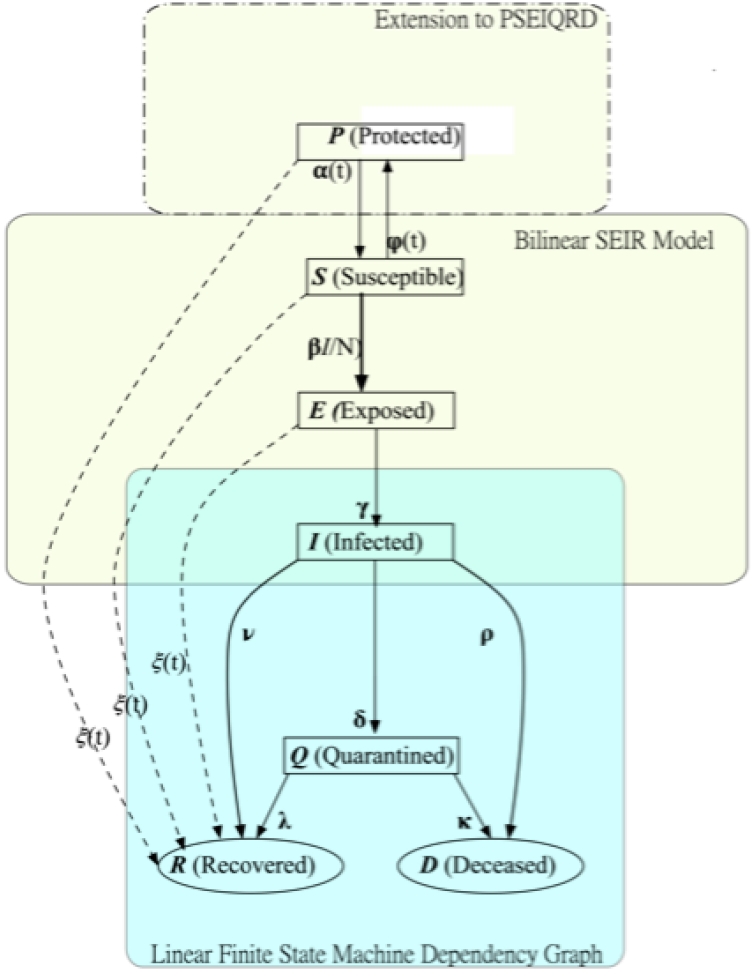
State Transition Graph of the Model

Note that the long dashed arrows are only exist in the presence of vaccination, as discussed below in Section 3.

This model is *conservative*, in the sense that it assures the total population is constant. That is,

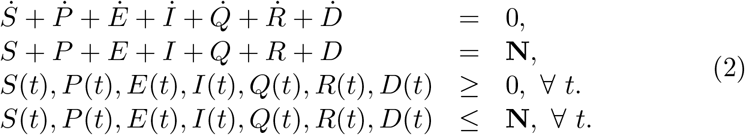

### 1.3 The Timed Interventions of the Model

We define 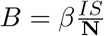 as the bilinear term of Eq. 1. The quasi-stable dynamical system defined by these equations must eventually reach *equilibrium* in which 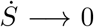 an *S*(*t*) → *S*_*F*_, and similarly for *P, E, I, Q, R* and *D*. Thus we can succinctly state definitions for 2 types of timed interventions at time *t* = *τ* :

1. Protection–if *αS*(*τ*) *> ϕP* (*τ*), then 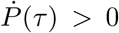 and population flows from *S* to *P*;
2. Release–if *αS*(*τ*) < *ϕP* (*τ*), then 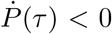 and population flows to *S* From *P*;

Note that intervention types are defined by the state of the system at the time of intervention, that is, at the start of the associated interval. Although it often happens that these conditions persist throughout the interval, sometimes they do not, due to the influence of the bilinear term *B*. The role played by *B* is crucial, and shall be closely monitored here.

The primary objective of our research is define, seek, and realize (theoretical) *virus extinction*–Suppose there exists *τ*_Ω_ such that *∀t > τ*_Ω_, *αS*_*F*_ = *ϕP*_*F*_ = *E*_*F*_ = *I*_*F*_ = *Q*_*F*_ = 0, but *R*_*F*_ and *D*_*F*_ are positive constants. In this case, since *E*_*F*_ = *I*_*F*_ = *Q*_*F*_ = 0 and *R*_*F*_ *>* 0, the virus, but not the host, is extinguished, and the pandemic is over.

Model parameter values have been shown to exist which demonstrate the existence of viral extinction.

Fortunately the condition that *E*_*F*_ = *I*_*F*_ = *Q*_*F*_ = 0 is not a rare case but is a guaranteed property of any equilibrium state of the model, as we now show. Here we use the definition 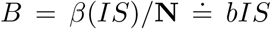, and define *ϵ = αP* − *ϕP*.

#### Theorem 1

*For any set of inputs to the dynamical system of Equation 1, (i) B*_*F*_ = *E*_*F*_ = *I*_*F*_ = *Q*_*F*_ = 0, *(ii), αS*_*F*_ = *ϕP*_*F*_ *>* 0, *(iii) E, I, and Q in general exhibit bell curve waveforms, and (iv) R and D exhibit logistic waveforms*.

**proof:** By Equation 1 and the above definitions, 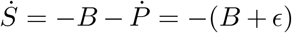, where 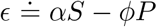. Similarly 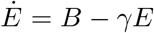. When equilibrium is reached, (denoted by *F* subscripts) we have 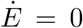, so *γE*_*F*_ = *B*_*F*_ = *bI*_*F*_ *S*_*F*_. In equilibrium 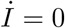 as well, so *γE*_*F*_ = (*δ* + *ρ* + *ν*)*I*_*F*_ and similarly *αS*_*F*_ = *ϕP*_*F*_ and *ϵ*_*F*_ = 0. Also, in equilibrium 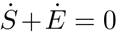, so *γE*_*F*_ = − *ϵ*_*F*_ = 0 and *E*_*F*_ = 0, and *I*_*F*_ = *γ/*(*δ* + *ρ* + *ν*)*E*_*F*_ = 0. Finally 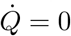 implies *Q*_*F*_ = (*δI*_*F*_)(*λ*+ *κ*) = 0 since *E, I* and *Q* start non-negative and asymptotically approach 0 for sufficiently large *t*, their shapes must resemble bell curves. Finally, since 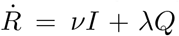, *R* is the integral of Gaussians and is hence logistic, and similarly for *D*.

#### Lemma 1

*Define µ*_*B*_ *to be the time of the final maximum of B*(*t*) *and similarly for µ*_*E*_, *µ*_*I*_, *an µ*_*Q*_. *Then*

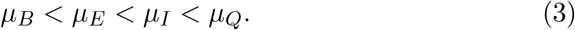

**proof:** Because in one day only a fraction of the population in a given state transfers to the other states, the complement of this fraction remains in the state (see Figure 2), the increase of *E* in the *i*_*th*_ time step which brought *E* to its maximum, cannot be propagated to *I* until at least the (*i* + 1)_*th*_ and possibly much later.

#### Theorem 2

*For any set of inputs to the dynamical system, there exists a set of interventions that lead to virus extinction*.

**proof:** Consider the function *B*(*t*) at time *t* = *µ*_*E*_.

Since 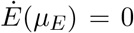, we have *B*(*µ*_*E*_) = *γE*(*µ*_*E*_) and since by the Lemma *µ*_*B*_ < *µ*_*E*_, it follows that *B* decreases to 0 asymptotically for *t > τ*_*E*_.

### 1.4 Interventions and COVID-19 in the USA Today

The real world per day waveforms of Fig. 1 are not Gaussian or logistic but appear to be piecewise combinations of such waveforms. Although there have been no formal governmental nationwide interventions, the data behaves as if there had been. In our experience, the pandemic appears to be lurching through a series of *punctuated equilibria*. Here we attempt to model these auto-interventions as closely as possible.

Note that in Figure 1, vertical lines have been drawn at 64^th^, 150^th^, 178^th^ and 250^th^ days. These are the days which we have found that changes in *α* and *ϕ* can generate the best match to data. The model attempts to capture these changes by defining the time dependence of the coefficients *α*(*t*) and *ϕ*(*t*) of Equation 1 to be piecewise constant. That is, the overall interval [0, *T*] is partitioned into *n* + 1 subintervals *i* = 0, 1, …, *n* with *T ≡ τ*_*n*+1_.

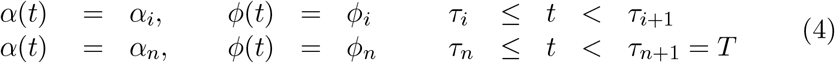

To understand *α* and the other population transition rate parameters, suppose that some time *t, S*(*t*) = 3*e*8. If *α* = 0.2, then at time *t*+Δ = *t*+1, if *B* and *P* happen to be negligible, *S*(*t*+1) = *S*(*t*) *–α* · *S*(*t*) *or* (3*e*8 − 0.45 *·* 3*e*8) = 1.35*e*8. In this case 135 million people are moved from *S* to *P* in a single (unit) time step of 1 day.^‡^

### 1.5 Assignable Parameters

Model definition is completed by the specification of the following assignable parameters:

1. a 9-vector *x* of assignable transition rate coefficients,where

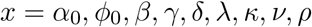

(see the state transition dependency graph of Figure 2);
2. *n* intervention time parameters *τ*_*i*_, *i* = 1, 2, …, *n*;
3. 2*n* assignable intervention rate parameters *α*_*i*_ and *ϕ*_*i*_, *i* = 1, 2, …, *n*.

Note that the initial rate parameters *α*_0_ and *ϕ*_0_ are the intervention rate parameters for the first interval. In general, there are 9 + 3*n* assignable parameters in all, not counting the 7 initial conditions on the state variables Each of these parameters must satisfy constraints on their allowable values shown below. The default values of these parameters (shown in square brackets) are the only set of transition parameters used to obtain the numerical results given here.

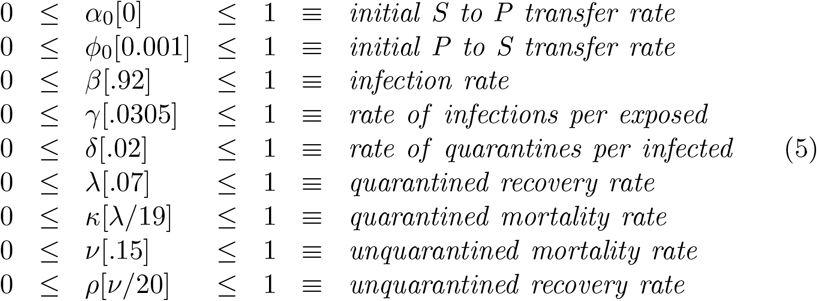

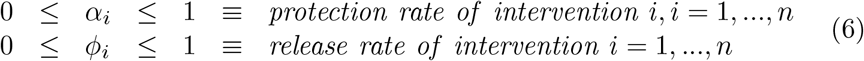

These define the constraints on the values the components of the *n*-vectors *α* and *ϕ*. A parameter set *p* ∈ *τ* × *x* × *α* × *ϕ* which satisfies the above constraints is called *feasible* and the solution of Equation 1 corresponding to such a *p* is called a *future* of the pandemic. A solution for an *infeasible* parameter set is a *behavior of the model* but not a *future of the pandemic*.

For the purpose of understanding the delays observed in the evolution of the pandemic, consider the node for the infected population *I*(*t*) in Figure 2. There are 3 possible state transitions out of this state, in which members of the population *I* are either:

1. quarantined with rate *δ*;
2. recovered with rate *ν*;
3. deceased with rate *κ*;

Of course some fraction of the infected population *I* do not transfer out but remain in *I*. Delays arise because it may take multiple, even many, days for increments of *I* arriving from *E* at any given time step to propagate forward. This argument applies to the non-terminal states *S, P, E*, and *Q* as well.

Numerical studies require the choice of the 7 initial conditions and the 9+ 3*n* free parameters. After first reviewing prior work on pandemic modeling, we shall compare the deaths and deaths per day data to date to the prediction of the model for the given rate coefficients and intervention parameters.

## 2 Summary of Prior Published Work on Pandemic Modeling

As stated above, our proposed model is an extension of that of Reference [2], which not only differs in the definitions of and 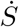 and 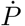, and of *I*, 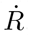, and 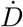, but also includes the *n* timed interventions with their associated 3*n* rate constants as well. Conceptually, our protected state *P* represents the isolated, or “sheltered in place” population rather than the innately immune. These differences enable the model to obtain excellent qualitative and quantitative results for the USA as a whole when compared to real world data to date.

Equation 4 and Equation 5 define 9+3*n* free parameters. The parameter space is 9 + 3*n*-dimensional (or 16 + 3*n* if we include the initial conditions for the 7-dimensional state space). The essence of modeling is to solve the *parameter identification problem*: Find the point in the 9 + 2*n* dimensional parameter space for which the model results best fit real world data. To solve this rigorously, a second round of research is proposed: find the optimal 2*n*+9 (2*n*+16) parameter assignments using the approach of Reference [10], starting from the quasi-optimal parameter set used throughout this paper.

Equation 1 specifies a set of 7 ordinary differential equations in which the 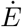 and 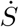 equations have a *bilinear* form which first appeared in Equation 29 of Reference [1]. More recently, they have been used in [11] to model a wide variety of infectious diseases. They can be reduced by parameter assignment to the Lorenz Equation [12], after the MIT professor who used them to model compartmental layers of the atmosphere. Such equations are known to exhibit unstable, and even chaotic, behavior, as treated in [13].

Fortunately, in our limited exploration of the multi-dimensional parameter space, no exotic behavior has been observed. Perhaps, the last 4 linear equations constitute a dissipative damping effect which inhibits the oscillatory behavior which might be generated by the state equations for *S* and *E*. In the sequel, it will be assumed that the system is stable.

Early epidemiological research [1] focused on detailed qualitative reasoning and Taylor series expansions. The lack of computing facilities did not stop them from considering a formulation even more general than that discussed here. They derived the SIR equations as a special case, and they found a special case solution in terms of hyperbolic functions for *z* = *R* + *D*. In this solution *ż* appeared to be Gaussian and *z* logistic, and they were able to assess viral extinction in their analysis.

Similar results from the same era were obtained in [6] whose author emphasized the same classification of population growth functions as logistic and Gaussian (except he used the term “derivative of logistic”). His solutions were consistent with Theorem 1 although no theorem was stated. Many more contemporary results, for example [7] and [5] had their own extensions to the SEIR model but the results could be similarly categorized.

None of these extended SEIR models achieved strong matches to COVID-19 data to date, because they were limited to one rise and fall of the bilinear Term *B*(*t*). With this limitation, the pandemic rises too rapidly, and then falls to extinction too rapidly to match data to date.

Statistical models [14] have achieved good short term matches, but do not consider extinction, which is a long term event. Such models cannot effectively forecast the duration of the pandemic. Recently, modeling works [4] have begun to appear that are sufficiently general to accommodate the influence of social and politcial changes in the midst of the pandemic, and have had some success predicting the winter surge the USA is currently experiencing. Reference [3] is worthy of note since they prove that any accurate analysis of the pandemic must be an extended SEIR model. Our work, also an extended SEIR model, extends it further by using the timed interventions. The necessity to treat the pandemic through multiple distinct epochs, in which the bilinear term rises, and falls as is clearly evidenced in the data to date. The timed-intervention SPEIQRD model permits direct projections of mortality and pandemic extinction, which is were clearly needed during the COVID-19 pandemic.

### 2.1 Extending Data Fitting Results into the Future with the Timed Interventions Model

Figure 3 illustrates the ability to use timed interventions to both tune the model to real data as well as perform long term predictions. In Fig. 3a we show the match between the model with timed interventions at *τ*_*i*_ = 64, 150, 178, 250 as compared to data, the corresponding values for *α*_*i*_ and *ϕ*_*i*_ are shown in Table 1. Here, by changing the flow of population between S and P we achieve an extremely good match to data extending all the way through the most recent date considered (Dec. 16^th^ 2020), validating the effectiveness of our model.

**Table 1:**
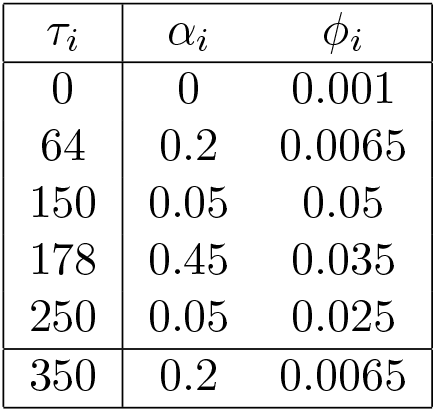
Values of Timed Intervention parameters for optimum match to data and predictions shown in Fig. 3

| $\tau_i$ | $\alpha_i$ | $\phi_i$ |
| --- | --- | --- |
| 0 | 0 | 0.001 |
| 64 | 0.2 | 0.0065 |
| 150 | 0.05 | 0.05 |
| 178 | 0.45 | 0.035 |
| 250 | 0.05 | 0.025 |
| 350 | 0.2 | 0.0065 |

**Figure 3:**
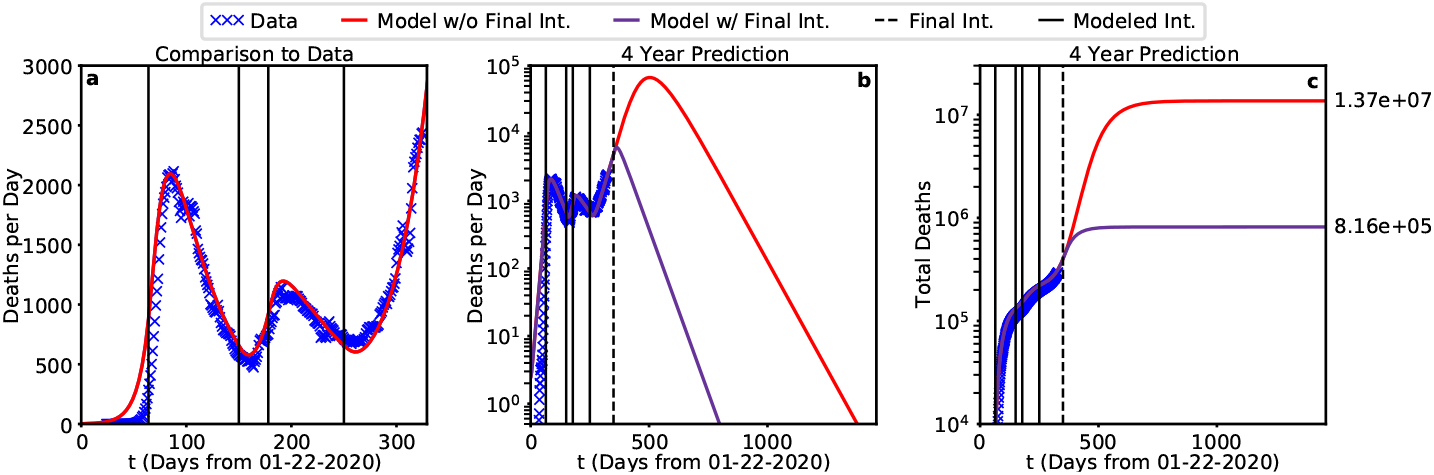
Best Fit to Deaths per Day and Total Deaths to Date.

We can then use the model to assess the influence of a potential fifth or final intervention at a date in the future. Figure 3b and 3c show the four-year predictions for Deaths per Day (b) and Total Deaths (c), where the modeled interventions from Fig. 3a are shown in solid lines with the new potential final intervention shown as a dashed line. Critically, we note that with a final intervention at day 350 (of comparable parameters to previous interventions) the deaths per day (purple curve) dips under 1 death per day in 766 days, an indication of viral extinction (see Section 3 for the definition). In contrast, without the final intervention (red curve) deaths per day doesn’t dip under 1 until the 1331^st^ day, more than 3.6 years out. Moreover, we can see the influence these two scenarios have on the total deaths that will occur during the pandemic. The scenario without a final intervention results in 17 times more deaths than with the final intervention, meaning that prompt action could (theoretically) save as many as 12.9 million lives.

A key focal point of this study has been forecasting the duration of the pandemic in the absence of a vaccine. Theorem 1 states that the waveforms for the size of exposed population *E*, infected population *I*, and quarantined population *Q* must asymptotically approach 0 as the duration of the simulation approaches infinity. Since the initial conditions for the size of these populations are positive, it follows that each of these sizes must reach a maximum value and then eventually decay to zero. The duration of the pandemic is, therefore, defined by how long It takes for these eventualities to occur. Since, by definition, 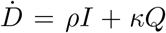 deaths per day must behave similarly. In fact, the red curve in Fig. 3b, beyond the 5^th^ intervention, strongly resembles a Gaussian function.

These eventualities are illustrated by the 2 year simulation of Figure 4. The red curves in the first column again indicate the absence of the 5t^th^ intervention. The purple curves illustrate the effect of the 5^th^ intervention and are seen to be significantly flattened by the this intervention. The days on which the red curves reach their maximum value are shown just above these maxima, which are ordered in accordance with Lemma 1.

**Figure 4:**
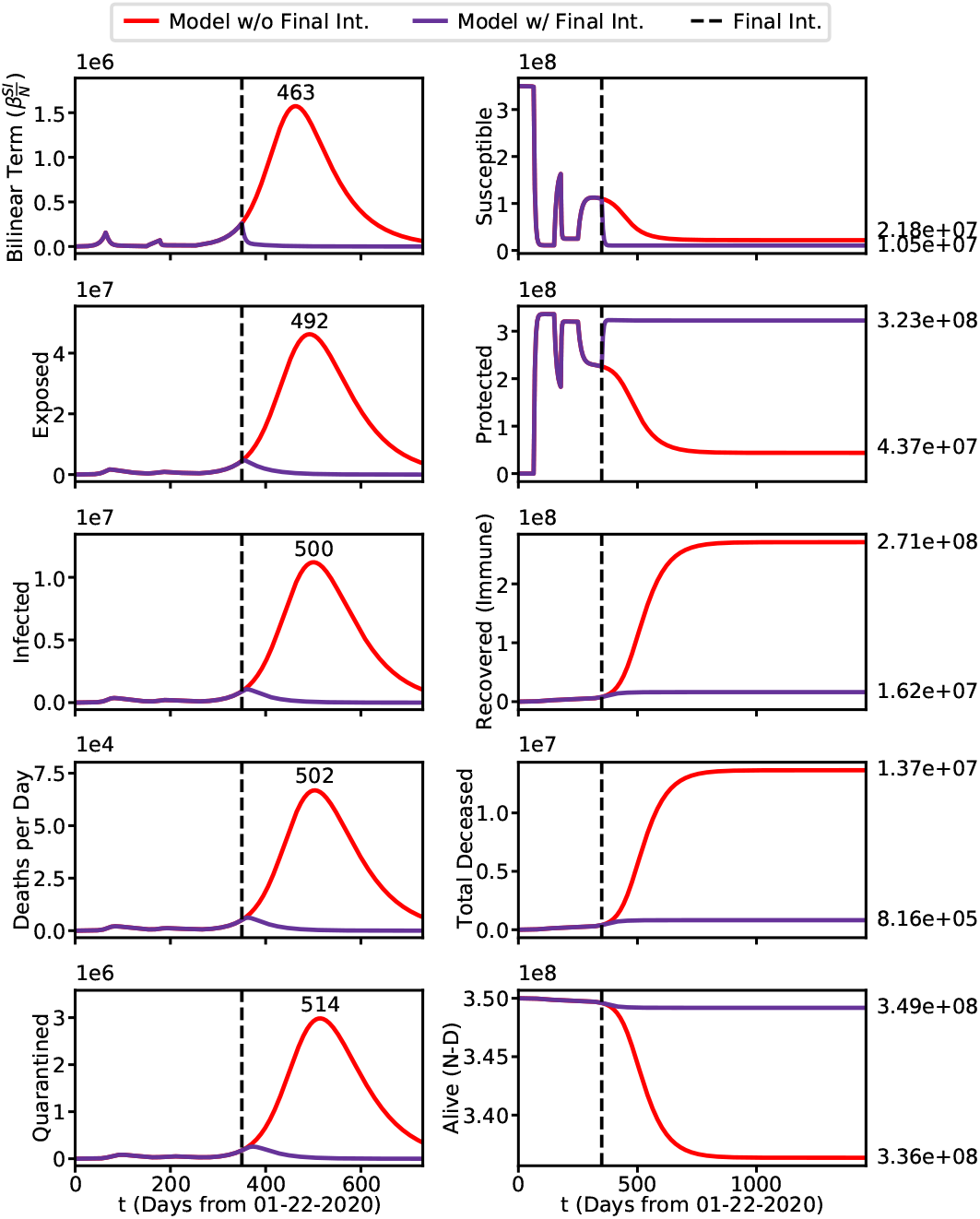
4 Year Predicted Waveforms–Best Fit Parameters.

On the right it is seen that in both scenarios *S* is drained by the last intervention (here extended out to a four year prediction). However, the red curve drains far less rapidly than the purple curve does in response to the strong protective 5^th^ intervention. However the Purple curve for *P* rises sharply in response to this intervention, whereas the red curve, instead of rising, is slowly drained at about the same rate as *S*. The purple *P* (*t*) curve reaches an asymptote just above 3e8, just beyond the 350^th^ day, the time of the 5^th^ intervention. In contrast, the red curve is tending toward an asymptote of 3e7, and has not reached its asymptote 2 years out. Note also that in the interval just before the intervention 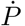 and 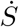 tend to approach 0 and according to Theorem 1, *αS*_*F*_ = *ϕP*_*F*_ at the end of that interval. We have called this effect “punctuated equilibria.”

Similarly the sink states *R* (labeled here as Recovered/Immune) and *D* approach constant values and display skewed logistic waveforms In accordance with Theorem 1. Again the gap between final death toll between the red and purple curves exceeds 1e7 (more than 10 million lives saved). Finally the bottom plot on the right illustrates *N* − *D*, a simple first definition of those that survive the Pandemic.

### 2.2 The Timing of the Final Intervention

As stated above, we have used theoretical interventions to determine a “best fit to data” model to simulate the behavior of the pandemic to date. During future prediction, however, we can employ future interventions, to maximize the number of survivors. These future interventions must be realizable in the political landscape of the United States. For example, in the “best case” future of Section 2.1 the critical last intervention increased the protected population to over 92% of the total U.S. population, and so is probably not politically feasible.

We now look at some alternative futures based on variations of the timing of the final intervention, shown in Figure 5. This is a complex figure but it can understood as follows. Each of the 16 plots has a red curve and purple curve. The red curves correspond to the absence of the final intervention, and therefore they do not change in any given column. Each of the 4 rows correspond to a specific time value for the final intervention, for *τ*_5_ = 350, 400, 450, 500, which is annotated with a dashed line. In each row the first waveform plotted is the bilinear term 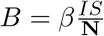. Looking down the first column, it can be seen that the maximum of *B* creeps up the red curve until the final intervention occurs and then decreases rapidly and quasi-logistically toward 0. The second column shows the effect of final intervention on the protected population *P*. It is seen that in each row the purple curve rapidly increases and stablizes quickly at a final value, but as the onset of the final intervention becomes later and later the total population that becomes protected gets lower and lower.

**Figure 5:**
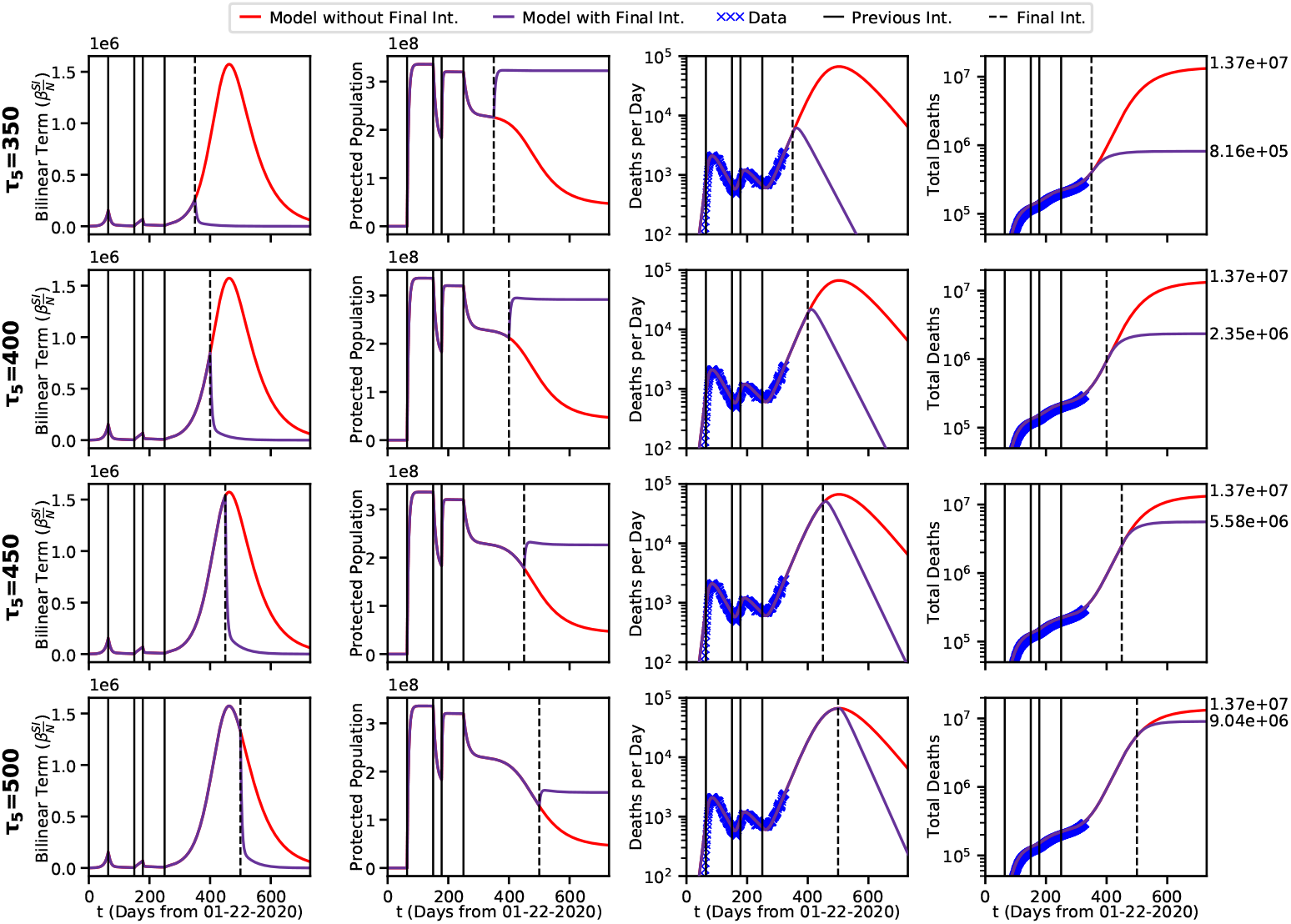
Four Possible Final Intervention Time Scenarios.

In the 3^rd^ column, we plot the deaths per day for each *τ*_5_. Here, we can see that this results in the date of viral extinction being pushed backwards significantly. This shows that even with another protective intervention if it does not occur soon viral extinction will not be achieved in 2021.

In the 4^th^ column it can be seen (purple curves) that the increase in the peak of the bilinear term incurs a corresponding increase of the corresponding final death toll, rising in the sequence 8.16e5, 2.35e5, 5.58e6, 9.04e6 as the day of the 5^th^ intervention increases in the sequence 350, 400, 450, 500. It is seen that the final death toll more than doubles for each 50 days of delay before the 5^th^ intervention. A similar result happens in the third column, where death per day reaches a maximum peak of around 5e4 in the bottom row.

It is also to be noted that with the exception of deaths per day, all of the protected scenarios show waveforms that have reached (or least come close to reaching) equilibrium within two years. However, in the *τ*_5_ = 500 scenario deaths per day does not go below 1 until day 924, nearly two years from the current date (dates where DPD goes below 1 shown for all scenarios in Table 2 below).

**Table 2:** Viral extinction dates and total fatalities as a function of day of final intervention, for the scenarios shown in Fig. 5

| $\tau_5$ | $\tau_{\Omega\dot{D}}$ | $\tau_{\Omega}$ | $D_F$ |
| --- | --- | --- | --- |
| 350 | 728 | 1047 | $8.16 \times 10^5$ |
| 400 | 829 | 1129 | $2.35 \times 10^6$ |
| 450 | 890 | 1164 | $5.58 \times 10^6$ |
| 500 | 924 | 1173 | $9.04 \times 10^6$ |
| None | 1331 | 1767 | $1.37 \times 10^7$ |

## 3 Vaccination Interventions

To account for the rollout of vaccination, we introduce a new coefficient to our model: *ξ*. This value represents the rate at which the population is immunized as a function of time (i.e. the rate at which population moves from *S, P*, and *E* to the recovered/immune state *R*. We can incorporate this into the model by treating vaccination as another timed intervention, where *ξ* is zero before the vaccine roll out. This corresponds to the following modifications of the basic model of Equation 1.

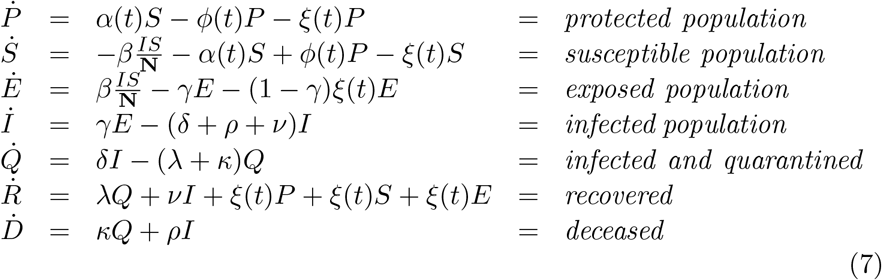

Note that each subtracted term is accompanied by an equal and opposite added term, so that the model still has the conservative property of Equation 1. The *ξ* terms correspond to the dashed arrows in Figure 2.

We use the same parameters as discussed above in the intervention case but only re-use the first 4 interventions. Thus the critical final protective intervention is replaced by a vaccination intervention of severity *ξ* that happens on the 350^th^ day. The modified model produces the waveforms illustrated in Figure 6. As before, the plots on the left show waveforms for the 5 Gaussian-like waveforms for the transient populations *B, E, I*, 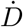, and *Q*. Here, the red curve is identical to the red curve in Fig. 4, which corresponds to no protective interventions or vaccinations, while the purple curve now represents the vaccination intervention.

**Figure 6:**
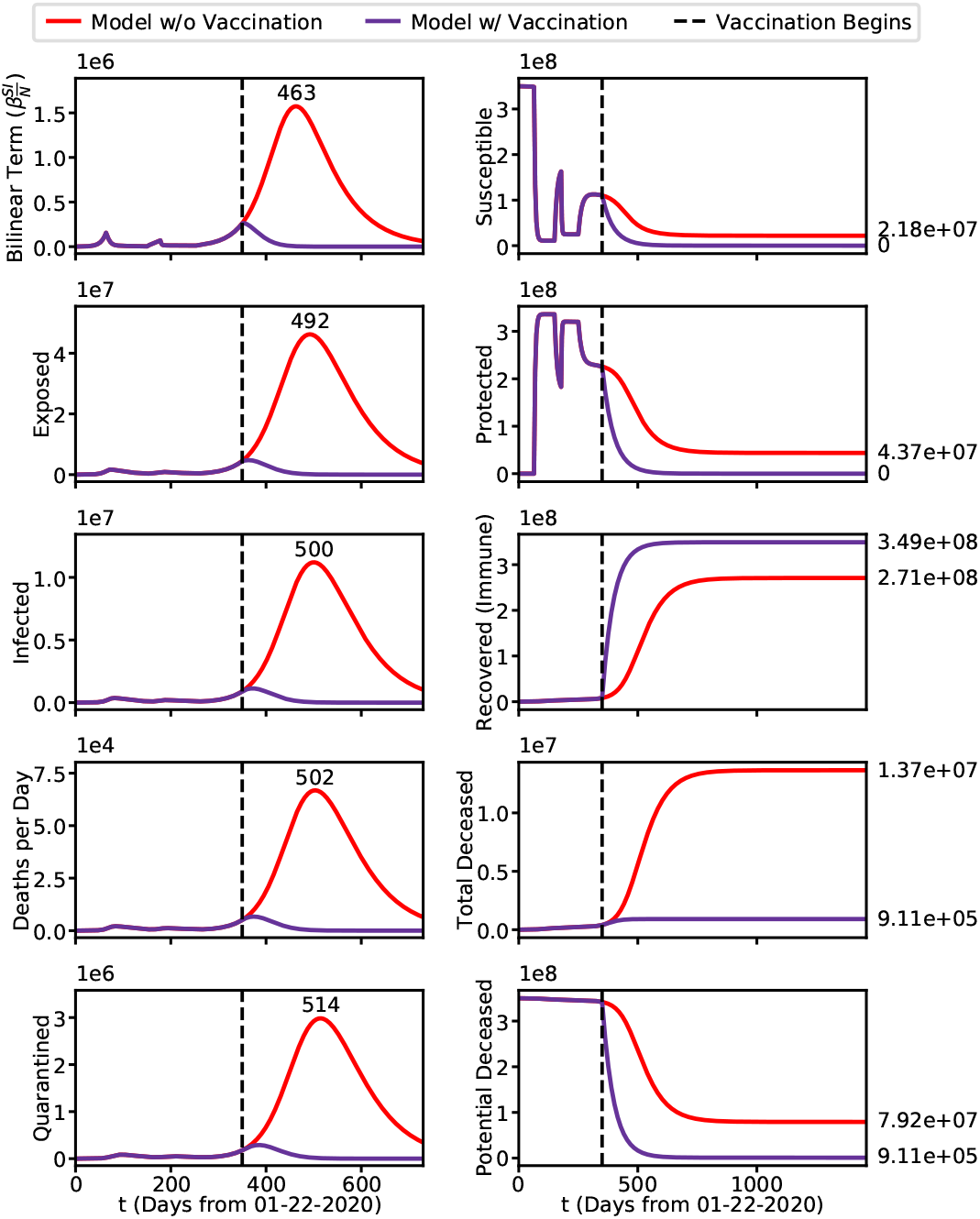
Vaccination Intervention Waveforms–Best Fit Parameters, with the final Protective Intervention replaced by a Vaccination Intervention of strength *ζ* = 0.02.

The plots on the right, however tell an entirely different story. The first two plots show that both purple and red curves are drained by vaccination in both plots the purple curves decay to 0, indicating the corresponding population are emptied. Without vaccination(red curves) the red curves reach equilibrium values of 2.18e7 (*S*) and 4.37e7 (*P*).

Note that the third plot is now labeled Recovered (Immune), rather than Recovered. The takeaway here is that the purple curve (3.49*e*8) is substantially above the red curve (2.71*e*8), and is close to the total population.

The fourth plot for the deceased is similar to the former case. However the bottom right plot is markedly different. There the ordinate is 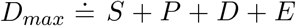 According to Theorem 1, in the final equilibrium we have *αS*_*F*_ = *ϕ*_*F*_ *P*, which given the specified final values of *α* and *ϕ* implies that *S*_*F*_ + *P*_*F*_ = 1.025*P*_*F*_, and since *P*_*F*_ is much smaller than before, and *E*_*F*_ = 0, it follows that *D*_*max*_ is much smaller than before. The text annotations at the right indicate that the red curve reaches an equilibrium value of 7.92e7 which is fully 87 times the equilibrium value 9.11e5 of the purple curve.

This is an important, possibly the most important, takeaway of our study: Whereas protective intervention can produce comparable mortality and viral extinction times, with vaccination the 3.49e8 people are immune, and cannot be reinfected.

As in the case of protective interventions, we summarize our vaccination results as shown in Figure 7.

**Figure 7:**
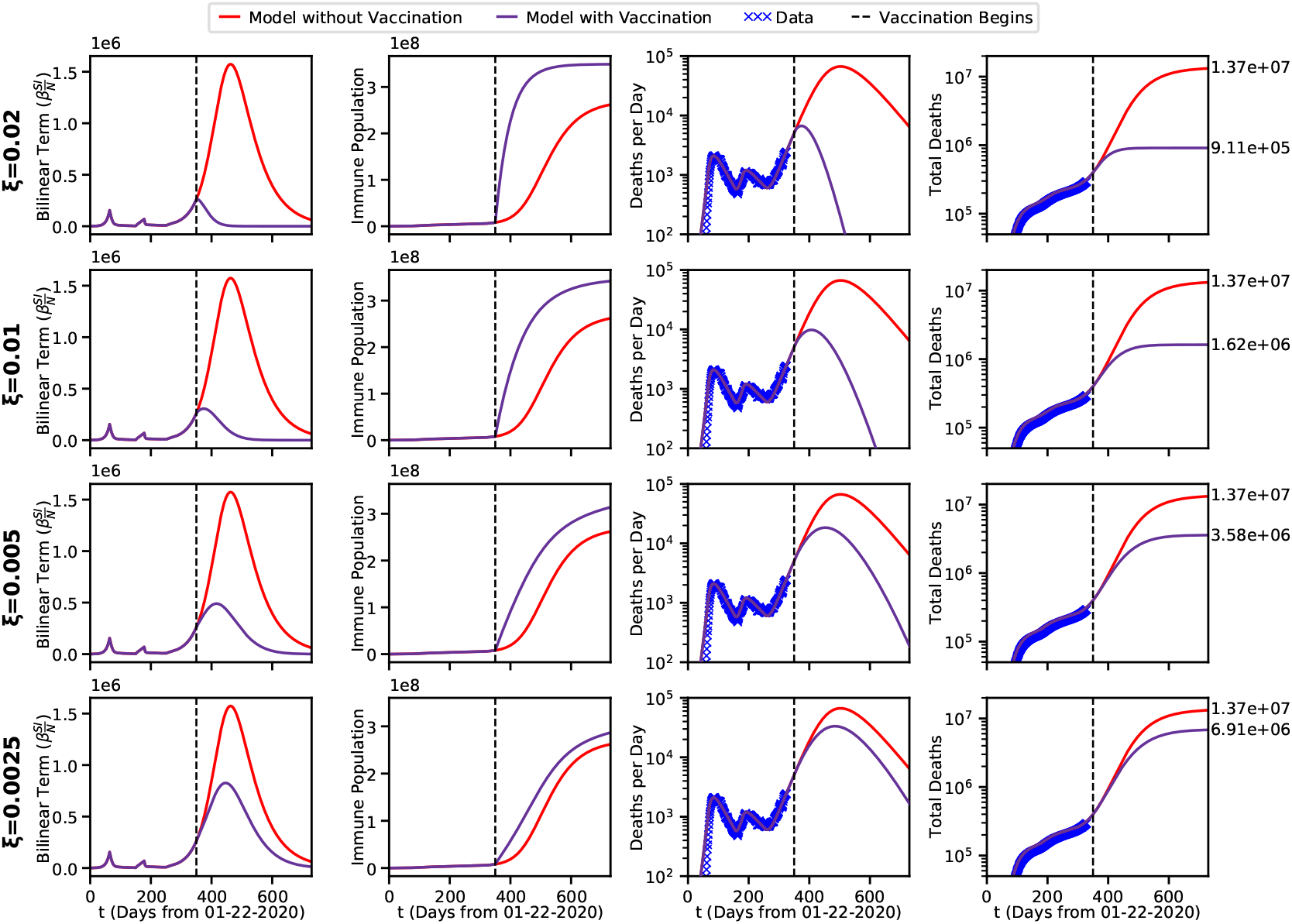
Four Possible Vaccination Strength Scenarios.

Looking down the first column, it can be seen that the maximum of *B* increases and creeps up the red curve as the strength of vaccination decreases. Arguably, the purple curve makes it to its 0-asymptote, at least in the first 3 rows. Clearly, the red curve doesn’t quite get there in the two year prediction shown. The second column shows the effect of the immuninization on the Recovered (Immune) population, *R*. It is seen that only the purple curve in the first row reaches quasi-equilibrium in 2 Years and only for the first row (Strongest Vaccination Intervention).

In the third column, the date of Viral Extinction can be determined from when deaths per day drops below 1, and the results are summarized below in Table 3 below.

**Table 3:**
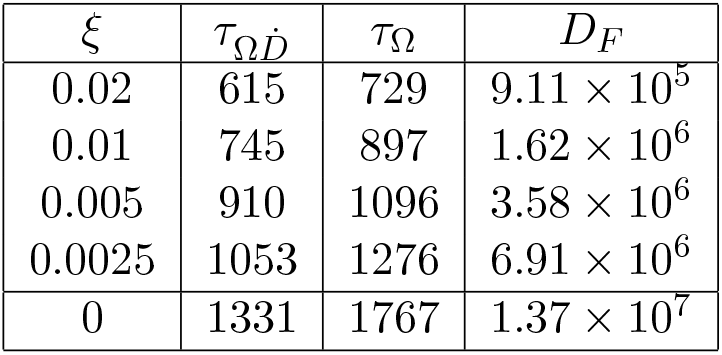
Viral extinction dates and total fatalities as a function of day of vaccination rapidity, for the scenarios shown in Fig. 7

**Table 4:** Initial conditions used for pandemic modeling

| $S_0$ | $P_0$ | $E_0$ | $I_0$ | $Q_0$ | $R_0$ | $D_0$ |
| --- | --- | --- | --- | --- | --- | --- |
| $N - (P_0 + E_0 + I_0 + Q_0 + R_0 + D_0)$ | 1 | 2000 | 500 | 0 | 0 | 0 |

The fourth column again corresponds to total Deaths.The text annotations at the right indicate total predicted Mortality with (red) and without (purple) vaccination of the strength indicated at the far left. Note the lives saved decreases from 12.8 million (*ξ* = 0.2) to 6.79 million (*ξ* = 0.0025).

Tables 2 and 3 summarizes the predictions of the proposed model on the Duration of the Pandemic. Here, we examine values concerning viral extinction. First, 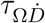 where 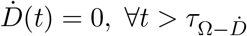 indicating that the worst of the pandemic is past, and true viral extinction, *τ*_Ω_, where *E*(*t*) = *I*(*t*) = *Q*(*t*) = 0, *∀t > τ*_Ω_ as defined in Section 1.3, as well as the total deaths.

Table 2 shows the effect of 5^th^ protective intervention without vaccination and Table 3 shows the effect of vaccination without the 5^th^ intervention. We see here, that for protective interventions the deaths can curbed and the worst of the pandemic can be avoided, but the pandemic must still run its course to achieve true viral extinction which will take multiple years. Moreover, since in the *P*_*F*_ is high for all the scenarios in Table 2 a further release intervention would result in another spike of the infected population and more deaths. Such a result would be consistent with the pandemic as observed in Europe where extensive social distancing mandates nearly eradicated the virus, but as those mandates laxed new surges were seen nearly everywhere. However, with extreme vaccination implemented immediately 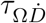 and *τ*_Ω_ can both be achieved within a year and tens of millions of lives can be saved.

## 4 Conclusions

A *timed intervention* extended SEIR model was presented and applied to the United States as a whole. In the discussion of Figure 4 and Figure 5. it was shown how, by controlling the magnitude of the bilinear term *B*, the *calamitous rapid rise and fall* of predator-prey models can be controlled, and even be actually be eliminated. Thus the desiderata of viral extinction can be realized. However it was also shown that this would require that around half of the total population be placed into lockdown which is unlikely to be feasible in the majority of high population density western first world countries.

These results were obtained with the most complete SEIR model extension employed to date, and with a parameter set which produces an almost perfect fit to the deaths per day and total deaths per day to date. (See the discussion of Figure 5.)

Our findings show that over a wide range of initial conditions and parameter sets, the presented model correctly predicts model behaviors with multiple qualitatively distinct phases. Also, we have given complete simulation and/or projected waveforms for all 7 of the component populations, whereas other studies focus more narrowly on vital statistics. We believe this model could be used, in conjunction with current economic factors, to recommend a direct intervention schedule which maximizes the number of survivors under realistic socioeconomic conditions.

We have presented extensive treatment of protective interventions of various degrees of timing values and severity. Also we have presented extensive treatment of the effects of vaccination. In this regard we have defined and computed viral extinction. For a representative set of supply dependent vaccination rollouts, we have given final mortality and theoretical final extinction dates.

A key finding of our study, is that unless prompt, effective protective interventions, or extensive vaccination interventions are introduced, the pandemic likely to extend all the way through 2021 and result in the deaths of many millions in the USA alone

The model is freely available for use and implemented in an easy-to-use iPython Jupyter Lab notebook (link in Methods section), meaning that epidemiologists, economists, and politicians can use the model immediately for help with long term predictions during the COVID-19 pandemic. This executable model could be immediately be used to examine smaller populations (i.e. individual states or counties) where one could quantify the mathematical influence of specific government interventions.

## 5 Methods

Data from this paper is taken from the COVID Tracking Project (www.covidtracking.com). The source is updated daily and all analyses presented in this manuscript are pulled directly from the API to present up to date values).

Final analyses were conducted on December 16^th^, 2020. The deaths per day data of Figure 1 were recorded by computing the consecutive day differences for the total death counts from the COVID Tracking Project. The thin line shows the real per day values, the thick line shows the seven day rolling average.

The ODEINT function from the python3 *SCIPY* distribution was used to solve these 7 ordinary ordinary differential equations over the closed time interval *t* ∈ [0, *T*], where *t* represents time in days, and *T* is the final time in days. Note that a one year (from January 22) simulation would only extend about 150 days into the future, the current time, *t*, is past the middle of the interval *t* ∈ [0, *T*]. We re-iterate the point that, in the simulations of this paper, only unit time steps are taken, as in displayed data from the model. Of course the odeint function takes arbitrarily smaller steps as required to deal with the waveform slope discontinuities frequently encountered in this study. Only the values at the unit times *t* = 1, 2, …, *T* are recorded. Fortunately, for the interesting part of the parameter space, the solutions of these equations are well-behaved, and have been used frequently as epidemiology models for over a century, including multiple times this millenium. Computationally, this simulation is easy, even trivial. Numerically, however, the simulation can be difficult, involving subtraction of large, almost equal terms, as well as discontinuities in the first derivative of the computed waveforms. For all predictions we use the following initial conditions vector,

The model is implemented in an iPython notebook that can be easily altered (to account for different intervention parameters) and used to model and predict the pandemic. It is freely available for download at https://github.com/hachteja/Timed-Interventions.

## Data Availability

All data is available in the Projections section of the Worldometer/USA website

## 6 Acknowlegements

The authors are indebted to Professor Janet Brandsma of Yale University for several crucial consultations which helped maintained virological credibility in the development and exposition of the timed intervention model. Thanks also to Professors Frank Stephenson Barnes and David Bortz of the University of Colorado, Boulder, who advanced our awareness of work in Engineering and Mathematical Biology. J.A.H. is supported by the Center for Nanophase Materials Sciences, which is a DOE Office of Science User Facility.

## 7 Author Contributions

G.D.H. conceived of the project, created the timed interventions model, and wrote the manuscript. J.D.S. guided with the direction of the project and the writing of the manuscript. J.A.H. wrote the companion iPython notebook and produced the figures. All authors participated in the revision of the manuscript.

## 9 Data Availability Statement

All data generated or analyzed during this study are included in this published article.

## Footnotes

† ”How long, I wondered, will this thing, last?? Lyric from “A Foggy Day” by George and Ira Gershwin

‡ We shall use the syntax 3e8 to denote 3 × 10^8^.

